# Birthweight, Childhood Obesity, Adulthood Obesity and Body Compositions, and Gastrointestinal Diseases: A Mendelian Randomization Study

**DOI:** 10.1101/2022.12.09.22283274

**Authors:** Shuai Yuan, Xixian Ruan, Yuhao Sun, Tian Fu, Jianhui Zhao, Minzi Deng, Jie Chen, Xue Li, Susanna C. Larsson

## Abstract

**Background:** Obesity has been established as a risk factor for several gastrointestinal diseases, whether the associations are causal is uncertain. In addition, the associations of obesity-related factors with gastrointestinal diseases have been scarcely explored. This Mendelian randomization aimed to investigate the associations of birth weight, childhood body mass index (BMI), adulthood BMI and waist-hip ratio, and body composition with the risk of 24 gastrointestinal diseases.

**Methods:** Independent genetic instruments associated with the exposures at the genome-wide significance level (*P*<5×10^−8^) were selected from corresponding large-scale genome-wide association studies. Summary-level data for gastrointestinal diseases were obtained from the UK Biobank and large consortia.

**Results:** Genetically predicted higher levels of birth weight was associated with a lower risk of gastroesophageal reflux. Genetically predicted higher childhood BMI was associated with an increased risk of duodenal ulcer, non-alcoholic fatty liver disease, and cholelithiasis. However, the associations did not persist after adjusting for genetically predicted adulthood BMI. Genetically predicted higher adulthood BMI and waist-hip ratio were associated with 19 and 17 gastrointestinal diseases, respectively. Genetically predicted greater visceral adiposity was associated with an increased risk of 18 gastrointestinal diseases. There were no strong associations between genetically predicted whole body fat and fat-free mass indices with gastrointestinal diseases.

**Conclusion:** This study suggests that greater adulthood adiposity, measured as either BMI, waist-hip ratio, or visceral adipose tissue, is causally associated with an increased risk of a broad range of gastrointestinal diseases.

## Introduction

Overweight and obesity affect approximately 60% of adults and 30% children that causes a global pandemic (1, 2). Gastrointestinal disease is also a globally prevalent health issue and causes a large disease burden worldwide (3). Population-based epidemiologic studies have found positive associations of adulthood obesity with the risk of many gastrointestinal diseases at different sites, including gastrointestinal reflux (4), peptic ulcer (5), diverticular disease (6), colorectal cancer (7), and non-alcoholic fatty liver disease (8), which may hint a pan-adversity of excessive fat accumulation on gastroenterological system. However, the associations of adulthood obesity with other common gastrointestinal diseases have been investigated in a few studies with inconsistent findings (9). Birth weight and childhood obesity have been associated with adulthood obesity (10, 11). Body compositions that cannot be precisely assessed by general adiposity indicators have been revealed to be differently associated with the risk of gastrointestinal diseases (12, 13). Nevertheless, limited data have been generated to examine the associations of birth weight, childhood obesity, and body composition with the risk of gastrointestinal diseases. In addition, observational studies are prone to be biased by reverse causation (i.e., weight loss attributed to gastrointestinal dysfunction before the diagnosis) and confounding. Whether the established associations between obesity and gastrointestinal diseases are causal remains uncertain.

Mendelian randomization (MR) is an epidemiological method that uses genetic variants as instrumental variables to infer the causality of an exposure-outcome association (14). Since genetic variants are randomly assorted at conception and cannot be modified by the onset of disease, MR investigation is less likely to be affected by confounding and reverse causality. Even though previous MR studies identified the associations of adulthood body mass index (BMI) and waist-hip ratio (WHR) with the risk of gastroesophageal reflux, esophageal cancer, gastric cancer, diverticular disease, non-alcoholic liver disease, liver cancer, cholelithiasis, and pancreatic cancer (15-18), the MR associations of adulthood obesity with other gastrointestinal diseases, like peptic ulcer, irritable bowel syndrome and pancreatitis, remains unestablished. In addition, the associations of other obesity-related traits with the risk of gastrointestinal disease have not been scarcely investigated. Here, we conducted an MR study to examine the associations of birth weight, childhood BMI, adulthood BMI, and WHR, and three adulthood body compositions (visceral adiposity, fat mass, and fat-free mass) with the risk of 24 gastrointestinal diseases.

## Methods

The study design is presented in **Figure 1**. This MR investigation was based on publicly available summary-level data of genome-wide association studies (GWASs), including the UK Biobank study and large consortia (**Table S1**). All studies have been approved by corresponding ethical boards of the relevant institutions and participants had given informed consent.

**Figure 1.**
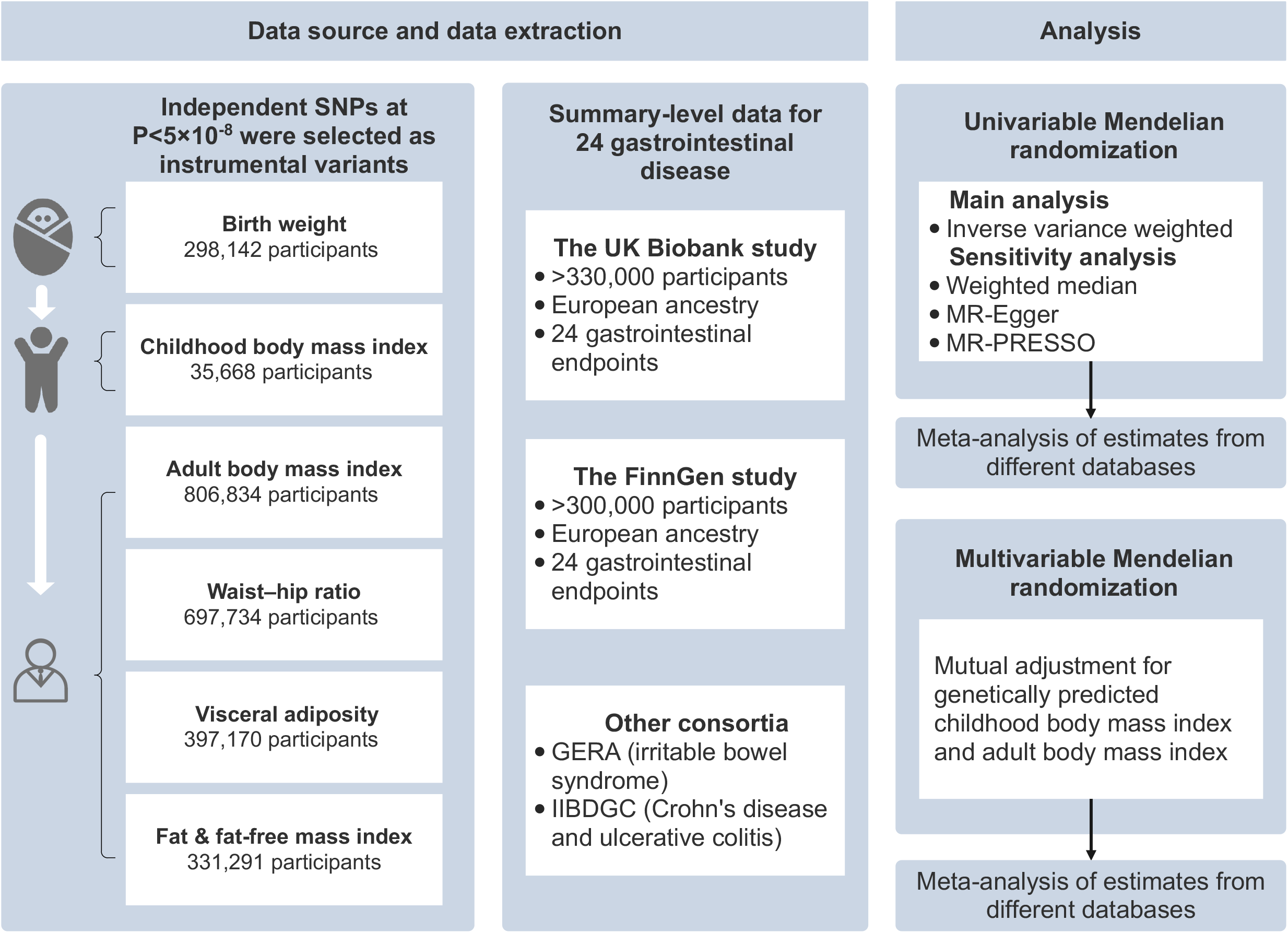
Study design. GERA, Genetic Epidemiology Research on Aging; IIBDGC, the International Inflammatory Bowel Disease Genetics Consortium; MR, Mendelian randomization; MR-PRESSO, Mendelian randomization pleiotropy residual sum and outlier; SNP, single nucleotide polymorphism.

### Genetic instrument selection

Genetic associations with birth weight and childhood BMI were extracted from the Early Growth Genetics Consortium (19, 20) with data in 298,142 European individuals for birthweight (19) and data in 35,668 European children aged 2-10 years for childhood BMI (20). For adulthood phenotypes, genetic associations with BMI and WHR were obtained from a GWAS of up to 806,834 individuals of European ancestries (21). The summary-level data on visceral adiposity were obtained from a GWAS in 396,220 European individuals, where visceral adiposity levels were estimated by a machine learning method with a training dataset of 4,198 European individuals with visceral adipose tissue measured by dual-energy X-ray absorptiometry (22). Fat-mass and fat-free mass were measured using bioelectrical impedance and corresponding genetic associations were obtained from the UK Biobank study including 331,291 individuals (23). Fat-mass index (FMI) and fat-free mass index (FFMI) were computed by dividing fat mass or fat-free mass by the square of height (23). We extracted single nucleotide polymorphisms (SNPs) associated with childhood BMI, adulthood BMI and WHR, visceral adiposity, and FMI and FFMI at the genome-wide significance level (*P* < 5×10^−8^) from the above GWASs. The linkage disequilibrium of selected SNPs for each trait was estimated based on the 1000 Genomes European reference panel. After removing SNPs in linkage disequilibrium (*r*^*2*^ ≥ 0.01), independent SNPs were used as instrumental variables in MR analysis. Detailed information on used genetic instruments is shown in **Table S2**.

### Outcome data sources

Summary-level data on 24 gastrointestinal diseases were obtained from the UK Biobank study (24), the FinnGen consortium (25), the International Inflammatory Bowel Disease Genetics Consortium (IIBDGC) (26), and Genetic Epidemiology Research on Aging (GERA) (27). The UK Biobank is a large population-based cohort study comprising over 500,000 individuals (24). In the UK Biobank study, summary-level statistics of the outcomes in the European ancestry were extracted from the GWAS conducted by the Lee lab where the gastrointestinal outcomes were defined using codes of the International Classification of Diseases 9th Revision (ICD-9) and ICD-10 (28). In FinnGen, summary-level data on gastrointestinal diseases were obtained from the R7 release (25). The FinnGen study is an ongoing nationwide study combining genetic and electronic health record data. Similarly, the gastrointestinal diseases in FinnGen were defined using the ICD-8, ICD-9, and ICD-10 codes. We additionally extracted summary-level genetic data on Crohn’s disease (5,956 cases and 14,927 controls) and ulcerative colitis (6,968 cases and 20,464 controls) from the IIBDGC, in which the patients were diagnosed by radiologic, endoscopic, and histopathologic evaluations (26). Data on irritable bowel syndrome (defined by the ICD-9 codes) were additionally obtained from the GERA, which included 3,117 cases and 53,520 controls (27). Detailed ICD codes used for outcome definition are shown in **Table S3**.

### Statistical analysis

The multiplicative random-effects inverse-variance weighted method was employed to calculate the primary causal MR estimate. Estimates for each gastrointestinal endpoint from different sources were combined using the fixed-effects meta-analysis. Heterogeneity among SNPs’ estimates was assessed by Cochran’s Q value. Three sensitivity analyses, including the weighted median, MR-Egger, and Mendelian randomization pleiotropy residual sum and outlier (MR-PRESSO) methods, were performed to detect horizontal pleiotropy and examine the robustness of the results. The weighted median method can provide accurate MR estimates when up to 50% of the instruments are valid (29). The MR-Egger intercept test can detect horizontal pleiotropic effect (30). MR-Egger can provide estimates albeit with low precision after adjusting for horizontal pleiotropy. MR-PRESSO method can identify horizontal pleiotropic outliers and provide estimates after the removal of the outliers (31). We performed multivariable MR analysis with adjustment for genetically predicted adulthood BMI for the identified associations between genetically predicted childhood BMI and gastrointestinal diseases to examine the independent role of childhood BMI. Multivariable MR was also conducted to estimate the associations of genetically predicted FMI and FFMI, which are strongly correlated traits, with gastrointestinal diseases. The Benjamini-Hochberg correction, which controls the false discovery rate (FDR) was applied to correct for the multiple testing separately for each exposure. The association with a *P*-value < 0.05 but Benjamini-Hochberg adjusted *P*-value > 0.05 were regarded suggestive, and the association with a Benjamini–Hochberg adjusted *P*-value < 0.05 were deemed significant. All analyses were performed using the TwoSampleMR and MendelianRandomization packages in R software (version 4.1.1).

## Results

### Birth weight

Genetically predicted higher levels of birth weight were associated with a lower risk of gastroesophageal reflux, duodenal ulcer, and non-alcoholic fatty liver disease (**Figure 2**). After correction for multiple comparisons, the association for gastroesophageal reflux persisted (OR [odds ratio] per standard deviation (SD) 0.87; 95% confidence interval [CI] 0.81-0.93) (**Figure 2 and Table S4**). This association was consistent in all sensitivity analyses (**Table S5**). There was evidence of heterogeneity between estimates of individual SNPs in the analysis of gastroesophageal reflux (**Table S5**). Pleiotropy was detected in the analysis of gastroesophageal reflux in the FinnGen study (MR-Egger intercept *P* <0.05), but the association remained in MR-PRESSO after removing the outlier (**Table S5**). Genetically predicted birth weight was not associated with the other studied gastrointestinal diseases (**Figure 2**).

**Figure 2.**
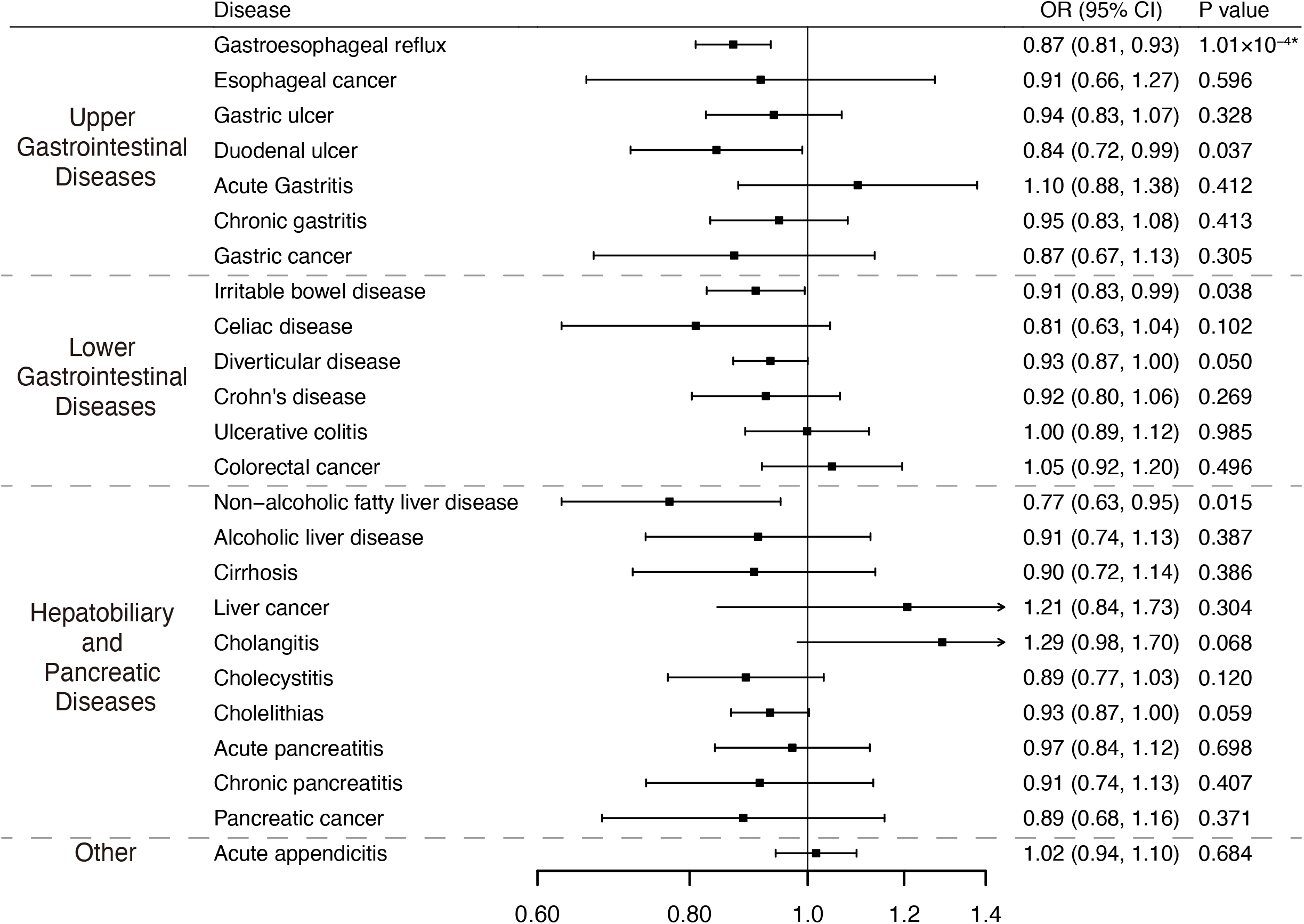
Associations of genetically predicted birth weight with 24 gastrointestinal diseases. CI, confidence interval; OR, odds ratio. * Significant association after multiple testing.

### Childhood BMI

Genetically predicted higher levels of childhood BMI were associated with an increased risk of esophageal cancer, duodenal ulcer, diverticular disease, non-alcoholic fatty liver disease, cholelithiasis, and pancreatic cancer (**Figure 3**). After correction for multiple comparisons, the OR was 1.31 (95% CI, 1.09-1.56) for duodenal ulcer, 1.39 (95% CI, 1.09-1.77) for non-alcoholic fatty liver disease, and 1.43 (95% CI, 1.30-1.57) for cholelithiasis per SD increase in genetically predicted childhood BMI (**Figure 3 and Table S4**). The associations remained directionally consistent in sensitivity analyses (**Table S6**).

**Figure 3.**
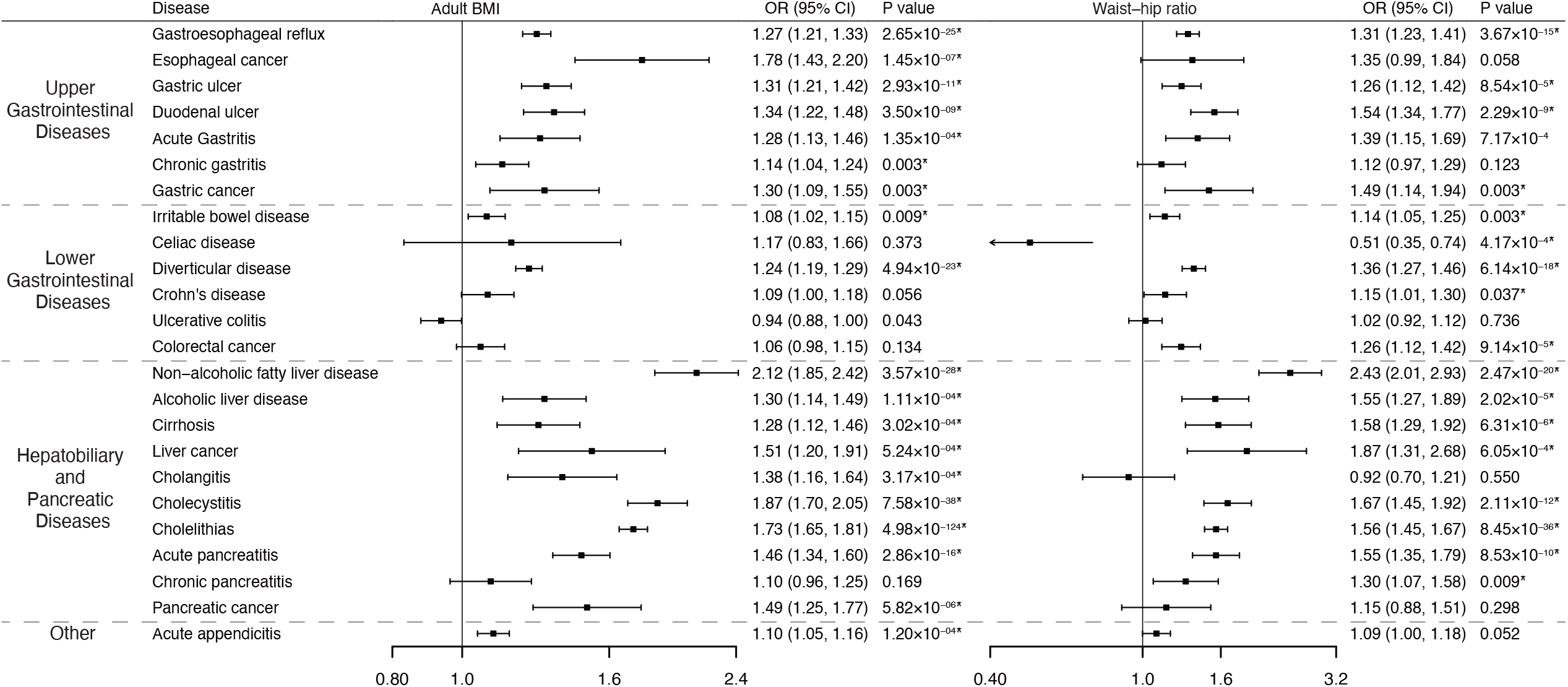
Associations of genetically predicted childhood body mass index with 24 gastrointestinal diseases. CI, confidence interval; OR, odds ratio. * Significant association after multiple testing.

Horizontal pleiotropy was observed in the analysis of cholelithiasis in UK Biobank (MR-Egger intercept *P* <0.05), but no outlier was detected in MR PRESSO (**Table S6**). Genetically predicted childhood BMI was not associated with other studied gastrointestinal diseases (**Figure 3**). The observed associations for genetically predicted childhood BMI attenuated and became nonsignificant in multivariable MR with adjustment for genetically predicted adulthood BMI (**Table S7**).

### Adulthood BMI and WHR

Genetically predicted higher levels of adulthood BMI were positively associated with 19 of 24 gastrointestinal diseases (**Figure 4**). All these associations remained significant after multiple-testing correction (**Table S4**). The OR per 1 kg/m^2^ increase in genetically predicted adulthood BMI was 1.27 (95% CI, 1.21-1.33) for gastroesophageal reflux, 1.78 (95% CI, 1.43-2.20) for esophageal cancer, 1.31 (95% CI, 1.21-1.42) for gastric ulcer, 1.34 (95%CI, 1.22-1.48) for duodenal ulcer, 1.28 (95%CI, 1.13-1.46) for acute gastritis, 1.14 (95%CI, 1.04-1.24) for chronic gastritis, 1.30 (95%CI, 1.09-1.55) for gastric cancer, 1.08 (95%CI, 1.02-1.15) for irritable bowel syndrome, 1.24 (95%CI, 1.19-1.29) for diverticular disease, 2.12 (95%CI, 1.85-2.42) for non-alcoholic fatty liver disease, 1.30 (95%CI, 1.14-1.49) for alcoholic liver disease, 1.28 (95%CI, 1.12-1.46) for cirrhosis, 1.51 (95%CI, 1.20-1.91) for liver cancer, 1.38 (95%CI, 1.16-1.64) for cholangitis, 1.87 (95%CI, 1.70-2.05) for cholecystitis, 1.73 (95%CI, 1.65-1.81) for cholelithiasis, 1.46 (95%CI, 1.34-1.60) for acute pancreatitis, 1.49 (95%CI, 1.25-1.77) for pancreatic cancer, and 1.10 (95% CI, 1.05-1.16) for acute appendicitis (**Figure 4**). Moderate to substantial heterogeneity was observed in the analyses; however, results were generally consistent in sensitivity analyses (**Table S8**). The MR-Egger intercept test indicated that there was pleiotropy in the analysis of gastroesophageal reflux, irritable bowel syndrome in UK Biobank, and diverticular disease (MR-Egger intercept *P* <0.05, **Table S8**). MR-PRESSO detected 1-14 outliers in these analyses, and the associations remained after the removal of the outliers (**Table S8**). There is a suggestive inverse association between genetically predicted adulthood BMI and the risk of ulcerative colitis (OR=0.94; 95% CI, 0.88-1.00) (**Figure 4 and Table S8**). Genetically predicted adulthood BMI was not associated with other studied gastrointestinal diseases (**Figure 4**).

**Figure 4.**
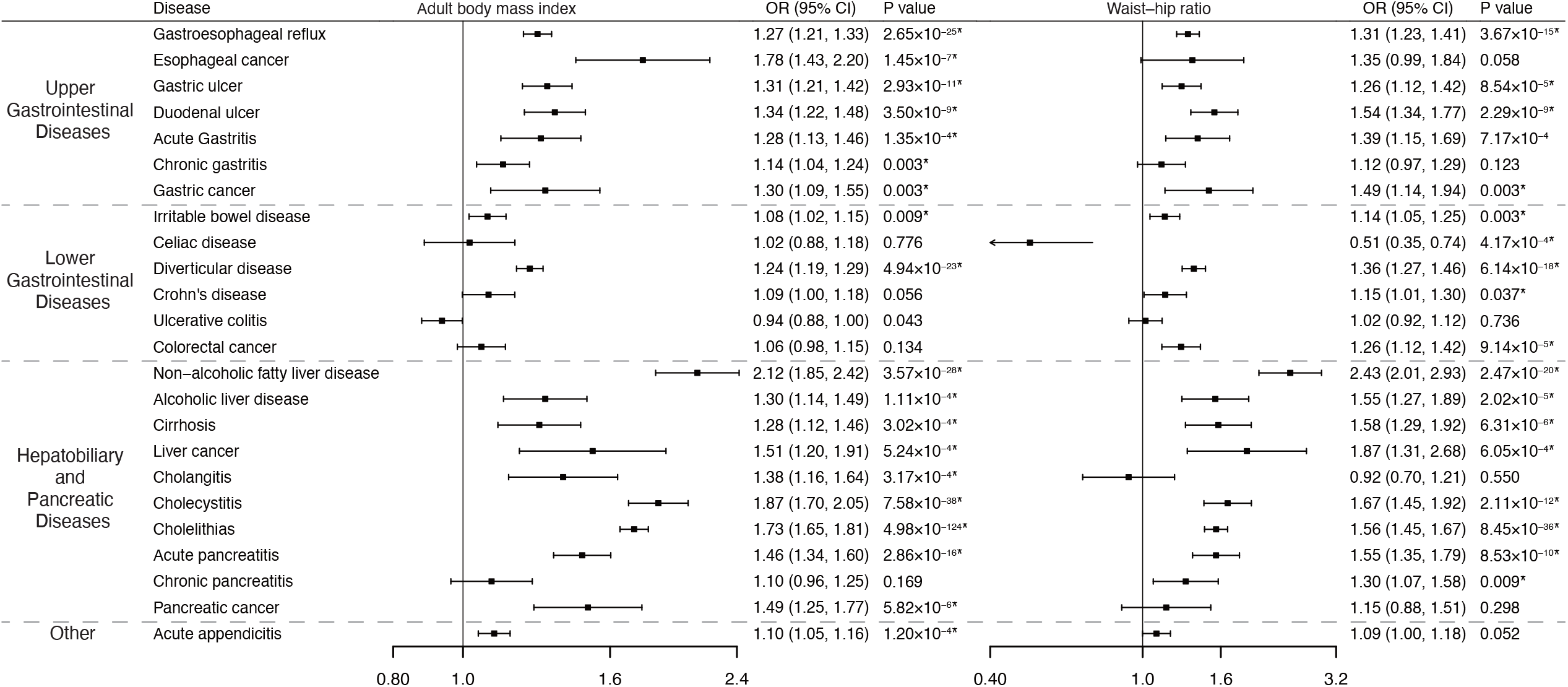
Associations of genetically predicted adulthood body mass index and waist-hip ratio with 24 gastrointestinal diseases. CI, confidence interval; OR, odds ratio. * Significant association after multiple testing.

Results for adulthood WHR were generally similar to that of adulthood BMI (**Figure 4**). Genetically predicted levels of WHR were associated with 17 gastrointestinal diseases after adjustment for multiple comparisons (**Figure 4 and Table S4**). Results were mostly consistent across sensitivity analyses (**Table S9**). Pleiotropy was detected in the analysis of gastroesophageal reflux and ulcerative colitis (MR-Egger intercept *P* <0.05); however, the results were persisted in MR-PRESSO after the removal of the outliers (**Table S9**). Genetically predicted adulthood WHR was not associated with other studied gastrointestinal diseases (**Figure 4**).

### Fat mass and fat-free mass indices

Genetically predicted higher levels of visceral adiposity were associated with 18 of 24 gastrointestinal diseases at the nominal significance level (**Figure 4 and Table S10**). After correction for multiple comparisons, the association per one kg increase in genetically predicted visceral adiposity persisted for gastroesophageal reflux (OR=1.34; 95% CI, 1.25-1.43), esophageal cancer (OR=1.43; 95% CI, 1.08-1.88), gastric ulcer (OR=1.26; 95% CI, 1.11-1.42), duodenal ulcer (OR=1.34; 95%CI, 1.22-1.48), acute gastritis (OR=1.33; 95%CI, 1.12-1.59), diverticular disease (OR=1.23; 95%CI, 1.15-1.31), non-alcoholic fatty liver disease (OR=2.45; 95%CI, 2.01-2.97), alcoholic liver disease (OR=1.38; 95%CI, 1.14-1.68), cirrhosis (OR=1.42; 95%CI, 1.18-1.70), liver cancer (OR=2.00; 95%CI, 1.43-2.81), cholangitis (OR=1.33; 95%CI, 1.04-1.70), cholecystitis (OR=1.62; 95%CI, 1.41-1.87), cholelithiasis (OR=1.77; 95%CI, 1.65-1.89), acute pancreatitis (OR=1.50; 95%CI, 1.31-1.71), chronic pancreatitis (OR=1.28; 95%CI, 1.07-1.53), pancreatic cancer (OR=1.84; 95%CI, 1.45-2.33), and acute appendicitis (OR=1.18; 95% CI, 1.10-1.26) (**Figure 4 and Table S10**). Results were overall consistent in the sensitivity analyses (**Table S10**). Moderate-to-high heterogeneity was observed in most analyses (**Table S10**). One to seven outliers were detected in MR PRESSO and all of these associations remained significant after the removal of the outliers. Genetically predicted visceral adiposity was not associated with other studied gastrointestinal diseases (**Figure 4**).

There were no associations of genetically predicted FMI or FFMI with gastrointestinal diseases after multiple testing corrections (**Table S4**). Genetically predicted higher levels of FMI were suggestively associated with an increased risk of duodenal ulcer, Crohn’s disease, and cholelithiasis (**Table S11**). Genetically predicted high levels of FFMI were suggestively associated with an increased risk of irritable bowel syndrome and a decreased risk of Crohn’s disease and ulcerative colitis (**Table S11**).

## Discussion

We conducted a comprehensive MR investigation to examine the associations of birth weight, childhood BMI, adulthood BMI and WHR, and three body composition measures (visceral adiposity, fat mass, and fat-free mass) with the risk of 24 gastrointestinal diseases. This MR study found that genetically predicted higher adulthood BMI, WHR, and visceral adiposity were associated with an increased risk of a broad range of gastrointestinal diseases (**Figure 5**). Genetically predicted childhood BMI was positively associated with some gastrointestinal diseases, but the associations appeared to be driven by adulthood BMI. No strong associations were observed for genetically predicted birth weight, FMI, or FFMI with any of studied gastrointestinal diseases.

**Figure 5.**
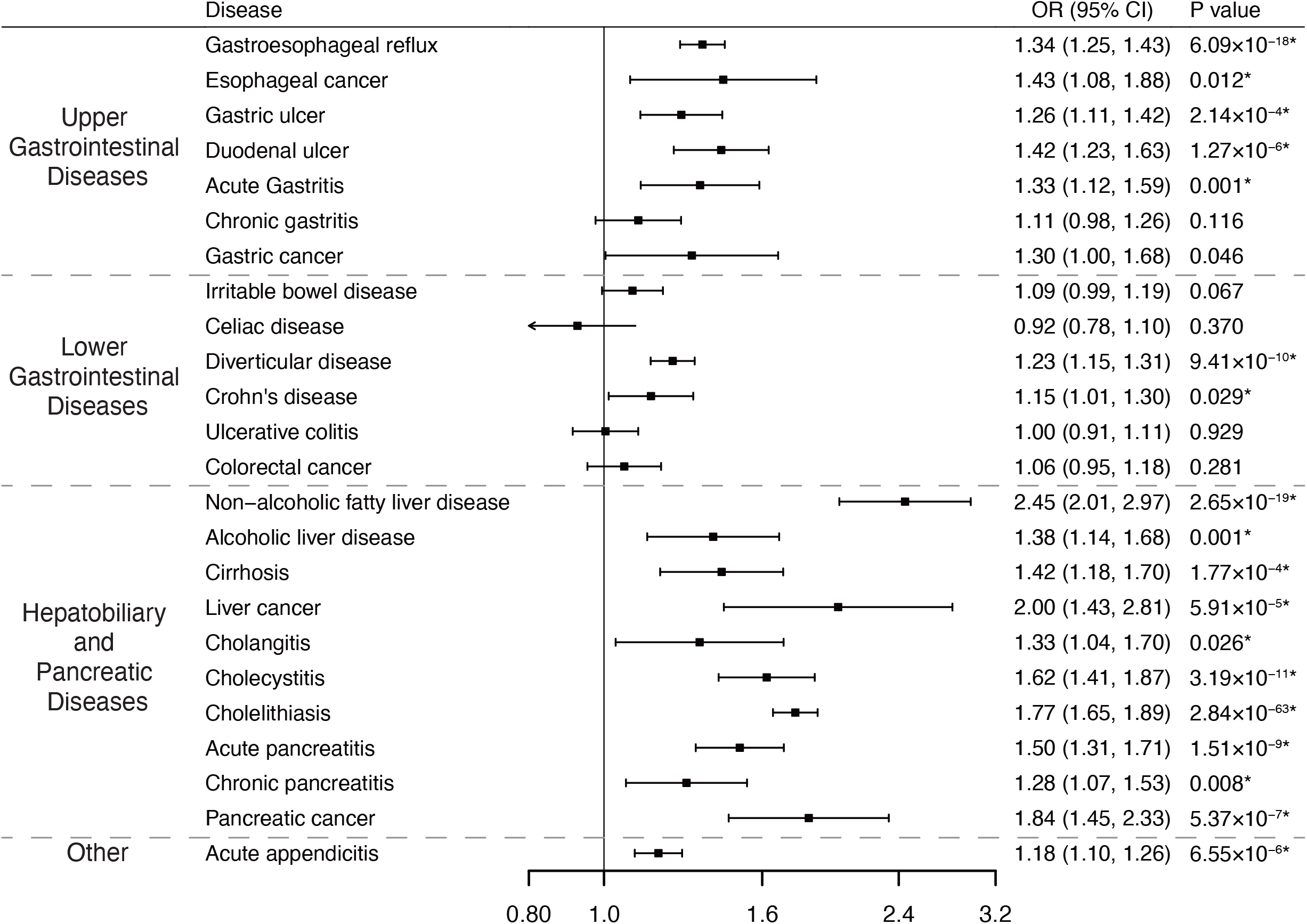
Associations of genetically predicted visceral adiposity with 24 gastrointestinal diseases. CI, confidence interval; OR, odds ratio. * Significant association after multiple testing.

**Figure 6.**
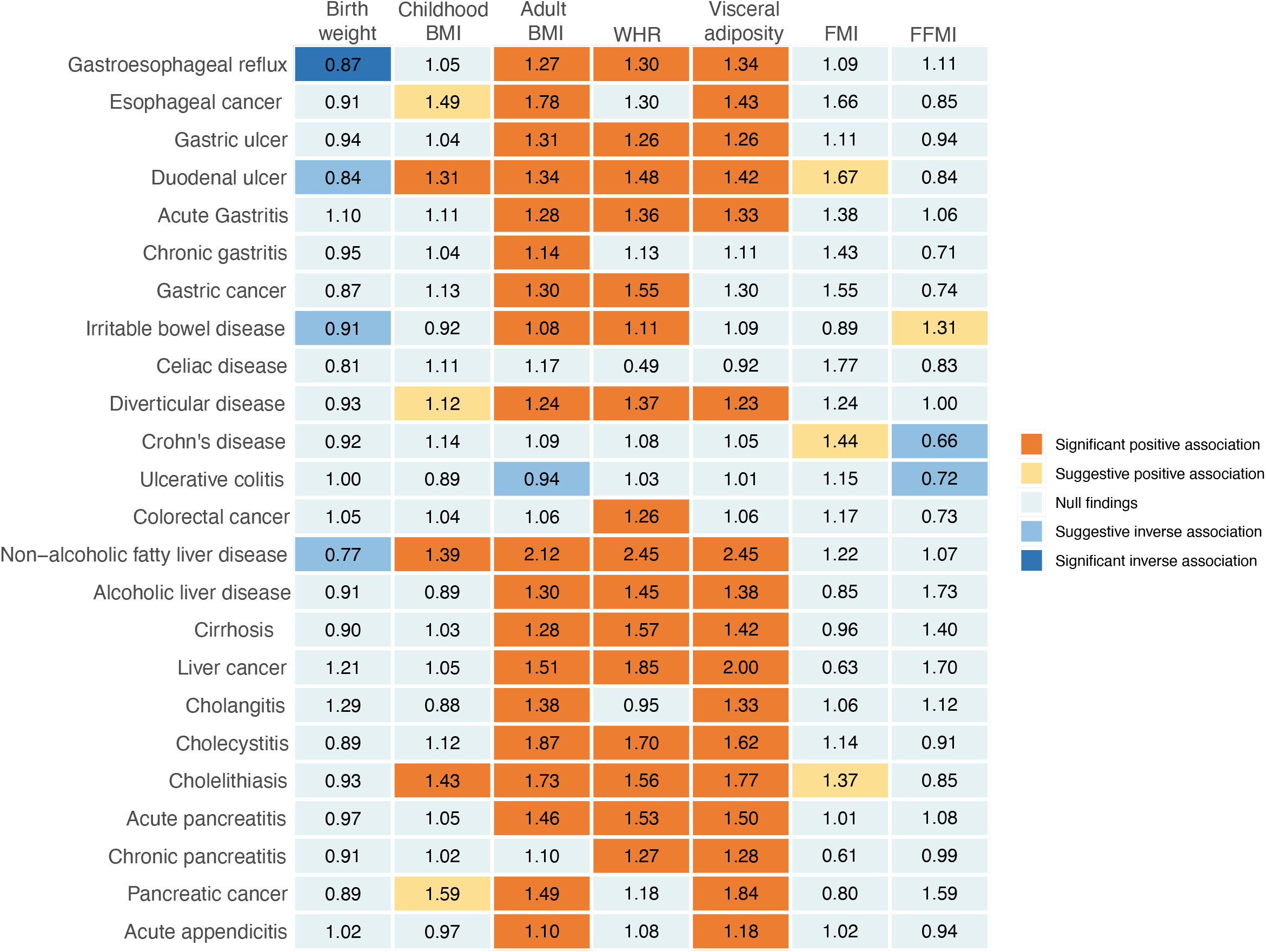
Summary of associations of genetically predicted birth weight, childhood obesity, adulthood obesity and body compositions with 24 gastrointestinal diseases. BMI, body mass index; WHR, waist-hip ratio; FMI, fat mass index; FFMI, fat-free mass index. The numbers in the box are the odds ratios of the associations. The association with a *P*-value<0.05 and Benjamini-Hochberg adjusted *P*-value>0.05 was regarded suggestive and the association with a Benjamini–Hochberg adjusted *P*-value<0.05 were deemed significant.

We found evidence that genetically predicted birth weight was inversely associated with the risk of gastroesophageal reflux, duodenal ulcer, and non-alcoholic fatty liver disease. Although the specific biological interpretation is unclear, several mechanisms might explain the associations. Birthweight reflects both intrauterine fetal growth and length of gestation. Reduced fetal growth rate may affect the relative development of organs, leading to persistent alterations in physiologic and metabolic homeostatic set points (32). Besides, the intrauterine fetal growth may cause changes in the state of the fetal genome and imprint gene expression and the epigenetic alterations in early embryos may carry over into subsequent developmental stages (33).

Studies on the associations between childhood obesity and the risk of subsequent adulthood gastrointestinal diseases are scarce. Our study for the first time, identified some positive casual associations of genetically predicted childhood obesity with duodenal ulcer, non-alcoholic fatty liver disease, and cholelithiasis. However, these associations did not persist after adjusting for genetically predicted adulthood obesity, which indicates that early-life obesity may elevate the risk of gastrointestinal diseases majorly by fat accumulation later in life. Reducing obesity in children is of importance to lower the risk of later-life gastrointestinal disease. The more important strategy for gastrointestinal disease prevention may be to break the pathway from childhood to adulthood obesity.

Previous observational studies explored the associations between obesity and gastrointestinal diseases. An umbrella review focused on gastrointestinal cancer found that excess body weight was positively associated with the risk of esophageal, gastric, colorectal, liver, and pancreatic cancer (34). In line with these studies, our results replicated previous MR findings that positive associations between BMI and the risk of gastric (35), esophageal (35), liver (15), and pancreatic cancer (36). However, the current MR investigation did not observe a positive association between genetically predicted BMI and colorectal cancer. Instead, we found a positive association between genetically predicted WHR and colorectal cancer. Previous observational studies also revealed obesity as a risk factor for several non-tumor gastrointestinal diseases, including gastrointestinal reflux (4), gastric and duodenal ulcer (5), acute and chronic gastritis (37), diverticular disease (6), non-alcoholic fatty liver disease (8), alcoholic liver disease (38), cirrhosis (8), cholecystitis (39), cholelithiasis (39) and acute pancreatitis (40), which is supported by the current study. In addition, we also observed the positive associations of genetically predicted BMI or WHR with irritable bowel syndrome, cholangitis, and acute appendicitis, which have not been well established and need verification.

BMI, as an indicator of adiposity, cannot precisely measure body composition. Visceral fat appears more harmful compared to other fat sites (41). Limited by the expensive and time-consuming technique, visceral adiposity is usually evaluated by hip circumference or WHR in observational studies. Corroborating and extending the previous observational studies, the current MR investigation demonstrated that genetically predicted visceral adiposity was associated with the risk of gastroesophageal reflux (42), esophageal cancer (43), diverticular disease (44), non-alcoholic fatty liver disease (45), cirrhosis (45), liver cancer (46), cholecystitis (47), cholelithiasis (48), acute and chronic pancreatitis (47), and pancreatic cancer (49). We additionally found that genetically predicted visceral adiposity was associated with an increased risk of gastric ulcer, duodenal ulcer, acute gastritis, alcoholic liver disease, and acute appendicitis, which is novel and needs verification. To our knowledge, this is the first MR investigation to assess the associations between genetically predicted visceral adiposity and gastrointestinal diseases.

The mechanisms underlying the associations between obesity and gastrointestinal disease have not been well understood. Except the obesity-related changes in anatomy, several cellular, and molecular factors may be involved in explaining these associations. A low-grade chronic inflammatory state is present in obesity, which was confirmed by the increased systemic levels of pro-inflammatory markers and cytokines released from adipose tissue (50). This inflammatory response is thought to depend on the activation of innate and acquired immune (51). Besides, insulin and insulin-like growth factor (IGF) signaling may also have a role in gastrointestinal diseases, especially gastrointestinal cancer. Clinical studies supported that plasma levels of insulin, and free IGF-1 were higher in obesity (52), which triggered an intracellular cascade and stimulated the proliferation of tumor cells (53). Moreover, there is a growing appreciation of gut microbiota in the promotion of obesity according to the different microbiomes between obese individuals and normal-weight individuals (54). High-fat diet altered the composition of gut microflora and thus increased the lipopolysaccharides levels, which causes chronic inflammation and insulin resistance (55).

Overall, our results reemphasize the impact of obesity on gastrointestinal health. Going further, we specified the visceral adiposity was a risk factor for a broad range of gastrointestinal diseases, which implied that excess visceral fat should receive adequate attention even in a normal BMI given that nonobese individuals may have an excess of abdominal fat (56). Greater public education and awareness of the association of obesity with gastrointestinal diseases are vital because obesity is a modifiable behavioral risk factor that could be prevented by lifestyle factor intervention.

The major strength of the present study is the MR design, which minimized confounding and reversed causality. Besides, this study with a larger sample size improved the statistical efficiency. Consistent results from different large summary sources supported the robustness of our findings and improved the precision of our findings. Some limitations are worthy of note. A major limitation of the MR study was horizontal pleiotropy; however, we conducted a series of sensitivity analyses that indicated limited pleiotropic effects. Even though we used several data sources, the number of cases for certain gastrointestinal diseases was still small, which means we might overlook some weak associations due to inadequate power. In addition, we could not evaluate nonlinear relationships based on summary-level data. We could not perform the sex-stratified analysis either. Furthermore, although the confinement to the European populations minimized population structure bias, our findings might not be generalizable to other populations with different genetic backgrounds. It is also worth noting that genetic variants reflect the impact of a lifelong difference in obesity, and thus we could not capture the weight fluctuations in the trajectory of life. An important limitation of the analysis of FMI and FFMI is the very low variance in these traits explained by the genetic instruments. This likely explains the lack of significant associations for FMI.

In conclusion, our MR investigation found that genetically predicted higher BMI, WHR, and visceral adiposity were associated with increased risk of a broad range of gastrointestinal diseases. These findings suggest that reducing obesity, particularly visceral adiposity, is an important strategy to lower the disease burden of gastrointestinal disorders.

## Data Availability

Data can be obtained upon a reasonable request to corresponding authors.

https://osf.io/qxhjc/?view_only=96a171ea8f9e45e3868bda8c31ecddbc

## Acknowledgement

We want to thank the Lee Lab, the FinnGen study, the International Inflammatory Bowel Disease Genetics Consortium (IIBDGC), and the Genetic Epidemiology Research on Aging (GERA) for sharing data.

## Data sharing statement

Data can be obtained upon a reasonable request to corresponding authors.

## Authors’ Contribution

All authors read and approved the final manuscript and author contributions statement using CRediT with degree of contribution:

Shuai Yuan (Conceptualization: Equal; Methodology: Equal; Formal analysis: Equal; Data curation: Equal; and Writing - review & editing: Equal)

Xixian Ruan (Conceptualization: Equal; Methodology: Equal; Formal analysis: Equal; Data curation: Equal; and Writing – original draft: Equal)

Yuhao Sun (Conceptualization: Supporting; Methodology: Supporting; Writing - review & editing: Supporting)

Tian Fu (Conceptualization: Supporting; Methodology: Supporting; Writing - review & editing: Supporting)

Jianhui Zhao (Conceptualization: Supporting; Writing - review & editing: Supporting) Minzi Deng (Conceptualization: Equal; Data curation: Equal; and Funding acquisition: Equal; Writing - review & editing: Equal)

Jie Chen (Conceptualization: Leading; Data curation: Equal; Writing - review & editing: Leading)

Xue Li (Conceptualization: Equal; Data curation: Equal; Funding acquisition: Leading; and Writing - review & editing: Leading)

Susanna C. Larsson (Conceptualization: Equal; Methodology: Equal; Data curation: Equal; and Writing - review & editing: Leading)

